# The impact of health-system pharmacists on hospitalizations in heart failure: a systematic review and meta-analysis

**DOI:** 10.1101/2024.10.17.24315714

**Authors:** Lorenz Van der Linden, Craig Beavers, Paul Forsyth, Christophe Vandenbriele, Ross T. Tsuyuki, Fatma Karapinar-Carkıt, Lucas Van Aelst

## Abstract

**Background:** Previous evidence has established the role of pharmacists in heart failure (HF) care. However, the specific role of health-system pharmacists within in- and outpatient settings for HF patients has been left unexplored. This systematic review and meta-analysis aimed to evaluate the impact of health-system pharmacy interventions on all-cause and HF hospitalizations.

**Methods:** A systematic literature search was performed using PUBMED and EMBASE, following PRISMA guidelines. Randomized controlled trials (RCTs) published up to May 2024 that evaluated the effects of health-system pharmacy interventions on hospitalizations in HF patients were included. The quality of the included RCTs was assessed using Cochrane’s risk-of-bias tool. Meta-analyses were performed using random-effects models, with odds ratios (OR) as summary measure. Heterogeneity was assessed using the I^2^ statistic and Cochrane’s Q test.

**Results:** In total, 11 RCTs involving 3576 patients were included in our review. The meta-analysis of 9 RCTs assessing all-cause hospitalizations (3472 patients, 927 events) demonstrated a significant reduction with pharmacist care (OR 0.67, 95% CI: 0.49–0.92, p=0.0119). The second meta-analysis of also 9 RCTs, focusing on HF hospitalizations (3442 patients, 504 events), showed similar results (OR 0.64, 95% CI: 0.48–0.87, p=0.0038). Heterogeneity was moderate for both meta-analyses. Sensitivity analyses confirmed the robustness of the results. Subgroup analyses indicated greater effectiveness in outpatient settings and for extended interventions.

**Conclusions:** Health-system pharmacist interventions significantly reduce both all-cause and HF-specific hospitalizations in HF patients. Our findings highlight the importance of integrating pharmacists into multidisciplinary teams to improve HF management for in- and outpatients (PROSPERO: CRD42024593583).

## Introduction

Heart failure (HF) has a major impact on patients and healthcare systems, with a prevalence of up to 2% in the developed world.^1^ It is particularly common among adults aged 70 years and older, affecting approximately 10% of this age group.^1,2^ While considerable heterogeneity exists between countries, the average age at diagnosis in Europe and Northern America is approximately 75 years.^3,4^ Poignantly, this older demographic represents the fastest-growing segment within the developed world. Despite a mostly stabilized incidence, the overall prevalence of HF continues to rise, partly due to the aging of the population.^2,5^

HF patients are frequently hospitalized. The numerous hospitalizations explain about two thirds of the incurred health costs, totaling up to 2% of national health budgets in the developed world.^6,7^ On average, a HF diagnosis carries a risk of one unplanned hospitalization each year.^1^ Clinically, HF remains a severe condition, with a 1-year mortality rate of about 10% among chronic outpatient HF patients.^5^ Prognosis is even more concerning after an acute HF hospitalization with 90-day outcomes including a 10-15% risk of mortality and a 20-30% risk of readmission.^8^ This does not take into account HF-related emergency department visits which do not lead to hospitalization, but also place a significant burden on healthcare systems and patients.

Several drug therapies are available to manage HF and improve clinical outcomes.^9,10^ Guideline-directed medical therapy (GDMT) in HF with reduced ejection fraction (HFrEF) has been shown to improve the overall quality of life, mitigate HF symptoms and reduce all-cause mortality as well as hospital (re)admissions. A recent network meta-analysis demonstrated that GDMT conferred a more than 50% risk reduction of all-cause mortality.^11^ Despite these benefits, GDMT uptake remains suboptimal which cannot be entirely explained by clinical factors such as hypotension, bradycardia, renal impairment or hyperkalemia.^12^ Clinical inertia has been identified as a major factor preventing patients from receiving the four-drug GDMT regimen or from achieving target doses.^13^

Various interventions have been proposed to improve GDMT uptake among different HF patient populations.^14^ Strategies include audit-and-feedback, electronic alerts or clinical decision support systems (CDSS), outreach programs, transitional care programs, nurse-led care, and multidisciplinary teams.^14,15^ Many of these interventions have proven successful in increasing GDMT uptake and improving clinical outcome in HF patients.^10^ However, their implementation in clinical practice remains insufficient. Health-system pharmacist interventions may be particularly effective in supporting healthcare teams to further improve clinical outcomes in HF, particularly hospital (re)admissions.^16^ Health-system pharmacists work in inpatient or outpatient settings and are often hospital-affiliated. In this review, community pharmacists and those providing home-based care are excluded based on this definition.

Prior meta-analyses by Koshman *et al*., Parajuli *et al*., Arunmanakul *et al*. and Schumacher *et al*. have examined the impact of pharmacist involvement on various HF outcomes.^17-20^ Koshman *et al*. conducted a review and analysis including RCT’s up to 2007, focusing on all-comer pharmacist interventions in HF.^17^ Parajuli *et al*. updated this analysis in 2019.^21^ In 2021, Arunmanakul *et al*. expanded the scope by incorporating team-based HF care, where pharmacists took on specific roles.^20^ However, this expansion made it less clear to what extent the interventions were primarily dependent on the pharmacists themselves, and the impact on HF hospitalizations was not reported. In the same year, the update by Schumacher *et al*. concentrated on outpatient, community and home-based pharmacy interventions. ^22^ Across these four meta-analyses, the nature of the pharmacist interventions and patient populations varied substantially. Interventions ranged from a single post-discharge telephone call to inpatient team interventions and longitudinal ambulatory care. Pharmacists delivered these interventions either independently or as part of a team-based approach, categorized aptly by Koshman *et al*. ‘pharmacist-directed’ and ‘-collaborative’ care, respectively.^17^

Importantly, HF inpatients, recently discharged patients and those with worsening heart failure, often seen in outpatient settings, are at the highest risk of (re)hospitalization.^8^ However, the abovementioned meta-analyses have not specifically examined the impact of health-system pharmacists on both all-cause and HF hospitalizations in these high-risk patients. Therefore, this study aimed to update prior meta-analyses by focusing on RCTs involving in- and outpatients with HF who received interventions from a health-system pharmacist.

## Methods

### Data sources

An evidence-based literature review was conducted according to the Preferred Reporting Items for Systematic Reviews and Meta-Analyses (PRISMA) guidelines for meta-analyses of interventional studies.^23^ The protocol was published online (PROSPERO: CRD42024593583). The data search was based on search terms previously reported by Parajuli *et al*., Schumacher *et al*. and Arunmanakul *et al*., which overlapped substantially.^18-20^ Our search focused on RCTs published in peer-reviewed journals, that investigated the impact of health-system pharmacy interventions on all-cause and/or HF hospitalizations. Studies were retrieved from the bibliographic databases PUBMED and EMBASE using the search terms: ‘heart failure’, ‘pharmacist’ and ‘randomized controlled trials’, which were adapted to the specific requirements of each database. Searches were limited to English language articles and covered the period from database inception to May 2024. Snowball sampling was used to identify additional publications for review. The full search queries for both PUBMED and EMBASE can be found in the Supplementary Appendix.

### Study selection

After removing duplicates, references were uploaded to a reference manager, and RCTs were reviewed for possible inclusion in the review and meta-analysis.^24^ Since most articles would likely overlap with previous meta-analyses – the most recent of which was published in 2021 with a search cutoff data of February 9, 2021 – we decided to have one reviewer (LVDL) initially screen all publications.^22^ This process was evaluated by another research pharmacist (CB). In case of any residual doubt, consensus was reached with a third research pharmacist (PF).

For each reference, titles and abstracts were screened, and relevant full-text articles were reviewed. Articles were included if they concerned primary RCT publications that provided data on hospitalizations and focused on HF patients exposed to a health-system pharmacy intervention. Health-system pharmacist interventions were defined as those provided in inpatient or outpatient settings, excluding community pharmacy interventions and home-based care. RCTs were excluded if HF was not present in all study participants or if the role of the pharmacist was not explicitly defined (i.e., if a study mentioned pharmacists as possible team members without clearly outlining their responsibilities).

### Data extraction

A data collection form was used to gather the following information from the included studies: authors, year, country, region and study population (sample size, ejection fraction (EF), age, setting of patient recruitment). Details on the pharmacy intervention were also recorded. Documented intervention components were categorized as medication reconciliation, medication review, promoting medication adherence, education (e.g., on HF or on non-pharmacological measures), and extended interventions (i.e., those extending beyond the hospital stay for inpatients or beyond the ambulatory clinic visit for outpatients). For each included RCT, data were extracted on the primary outcome, whether the study was powered for any outcome, whether hospitalizations were part of the primary outcome, and whether the study achieved significant results regarding its chosen primary outcome.

### Quality assessment

The quality of each RCT was assessed upon inclusion using the Cochrane Risk-of-Bias 2 (Rob2) tool.^25^ A traffic light plot was generated to visualize the quality of the RCT results.

### Outcome

We focused on all-cause as well as HF hospitalizations as reported in the included RCTs, acknowledging potential variations in the definition of HF hospitalizations across studies. Hospitalizations not only reflect the burden of disease and treatment efficacy but also represent a critical endpoint for healthcare resource utilization and patient management strategies.

### Analysis

All analysis were conducted in R with the RStudio interface, using the meta and bayesmeta packages. Given the anticipated variability in pharmacist interventions, a random-effects model was employed. The analysis was conducted with the odds ratio (OR) as the summary measure and the inverse variance method for pooling. Between-study heterogeneity (tau^2^) was estimated using the DerSimonian-Laird method. Data were visualized using forest plots and drapery plots.^26^ Heterogeneity was assessed with the I^2^ statistic and significance for Cochrane’s Q, considering I^2^>75% and/or p(Q)<0.05 as indicators of significant heterogeneous study results. The number-needed-to-treat (NNT) was estimated assuming a 13.1% 30-day unplanned readmission rate which was based on the recent work by Van Wilder et al.^27^

We generated funnel plots and performed Egger’s test to assess small study effects. To test the robustness of our findings, a sensitivity analysis was conducted by systematically excluding each study and re-running the meta-analysis as described above. Results of this analysis were summarized in a separate forest plot. The impact of study characteristics (setting, intervention components, sample size) was explored in separate subgroup analyses.

## Results

### Literature results

A total of 407 articles was retrieved and 65 of these were reviewed for inclusion. After excluding 54 studies, the final selection comprised 11 relevant RCTs, totaling 3576 patients (n_intervention_ = 1741, n_usual care_ = 1835). The median study sample was 134 (interquartile range (IQR) 73-201). The PRISMA flow can be consulted in Figure 1.

**Figure 1:**
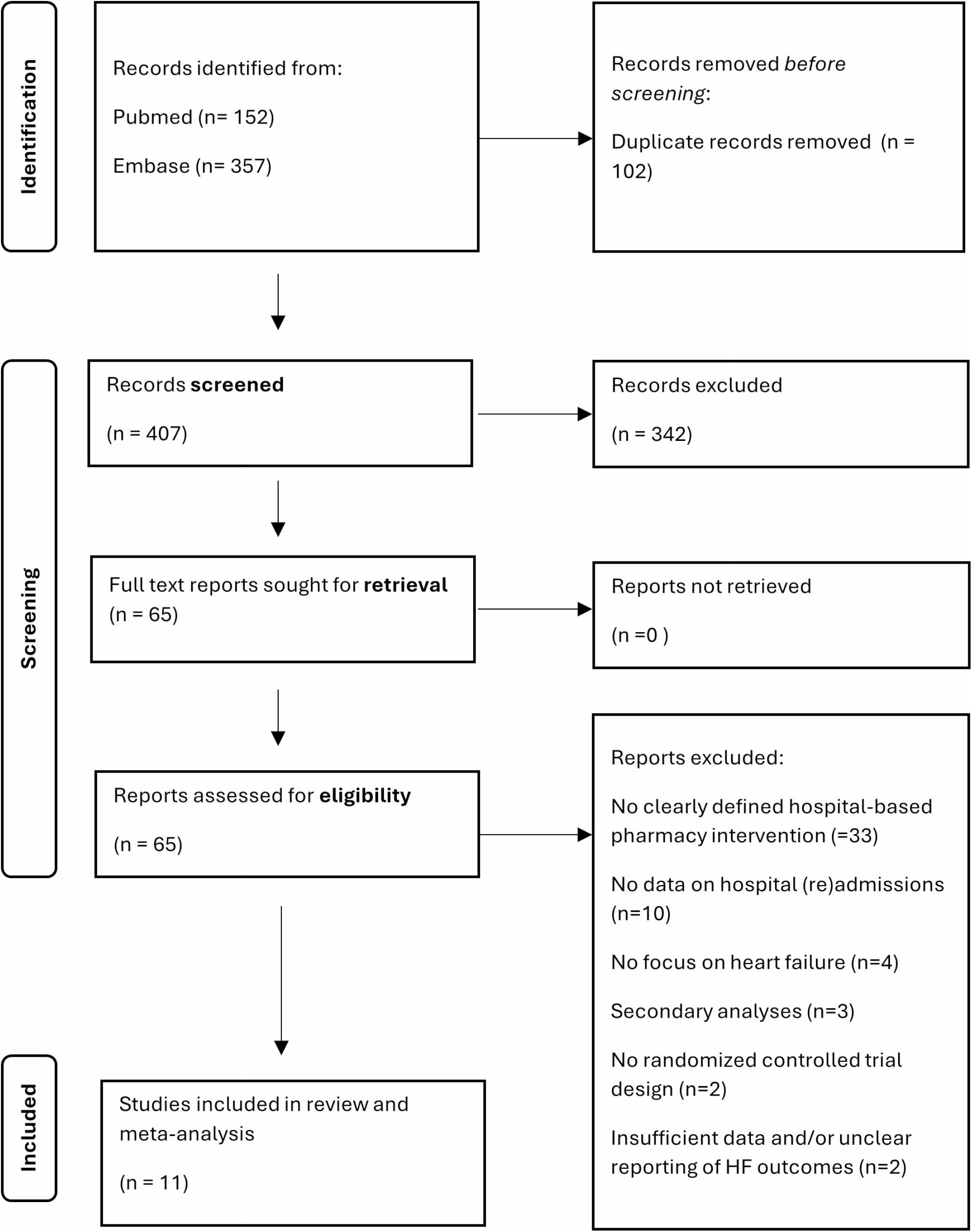
PRISMA flow diagram of study selection.

Included studies spanned between 1999 and 2020 and were conducted over various countries, eight of which were English speaking.^28-35^ Studies varied in their settings, with 3/11 focusing on outpatient care ^28,30,34^, 2/11 limited to inpatient care^32,36^, and 6/11 on both inpatient and outpatient care regarding patient recruitment and/or provision of the pharmacist intervention ^29,31,33,35,37,38^. Most RCTs provided medication reconciliation (n=8),^28-31,33,35,37,38^ medication reviews (n=7), ^28-31,35-37^ adherence promotion strategies (n=8),^28-33,37,38^ and patient education (n=9)^28-34,37,38^ Extended interventions, i.e., beyond the single contact moment, were provided in 8 RCTs.^28-31,33,35,37,38^ Five RCTs provided all five predefined intervention components.^28-31,37^ All studies, except for one, implemented the pharmacy intervention within a team setting.^38^ Sample size estimates were provided in 7 trials,^31,33-38^ 2 of which were powered for a clinical outcome, that included hospitalizations^35,38^. The median duration of follow-up was 12 months (IQR 6-12). Study characteristics can be retrieved in Table 1. Additional information on pharmacist interventions can be retrieved in the Supplementary Appendix (Table S1).

**Table 1:**
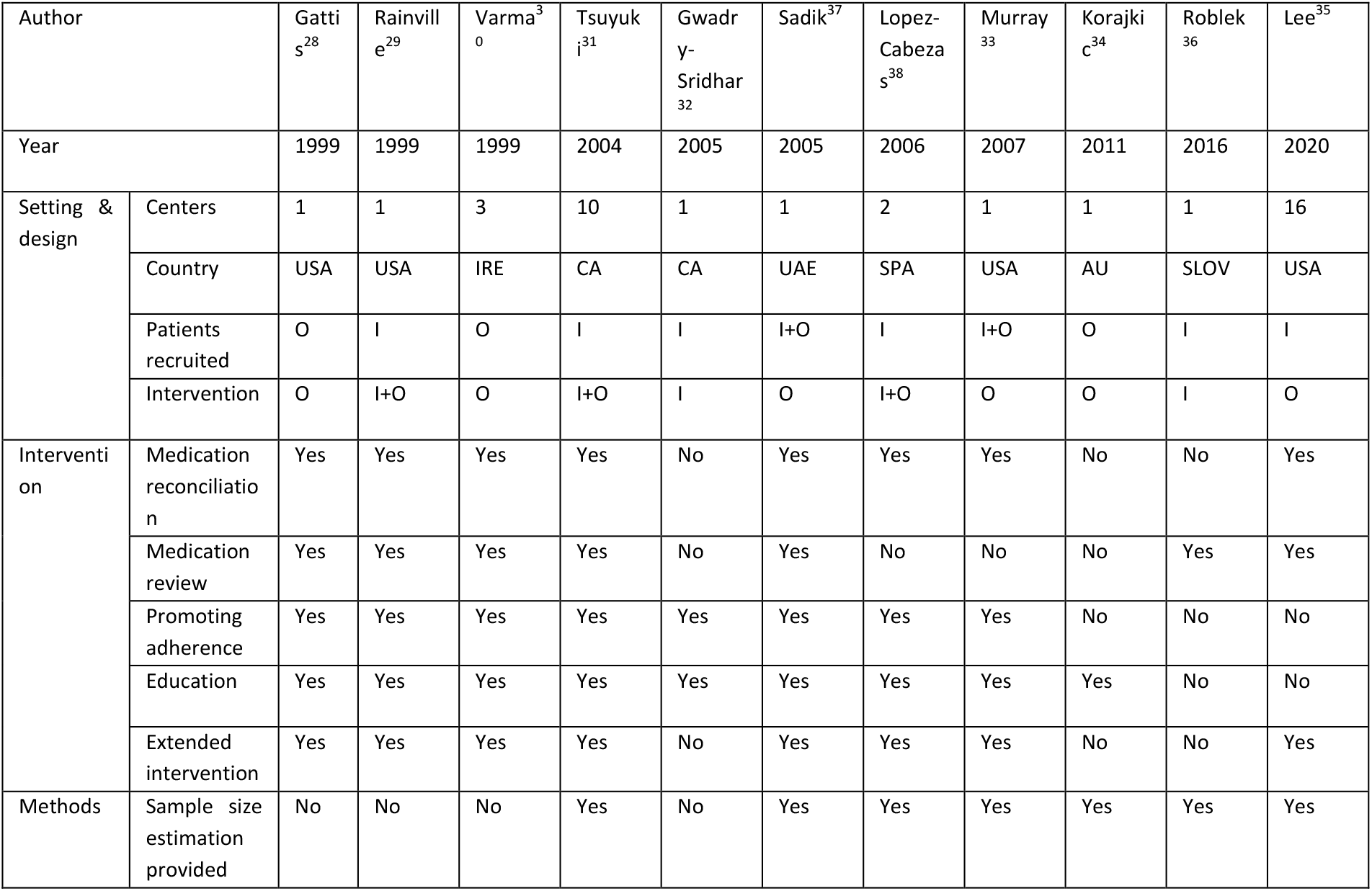
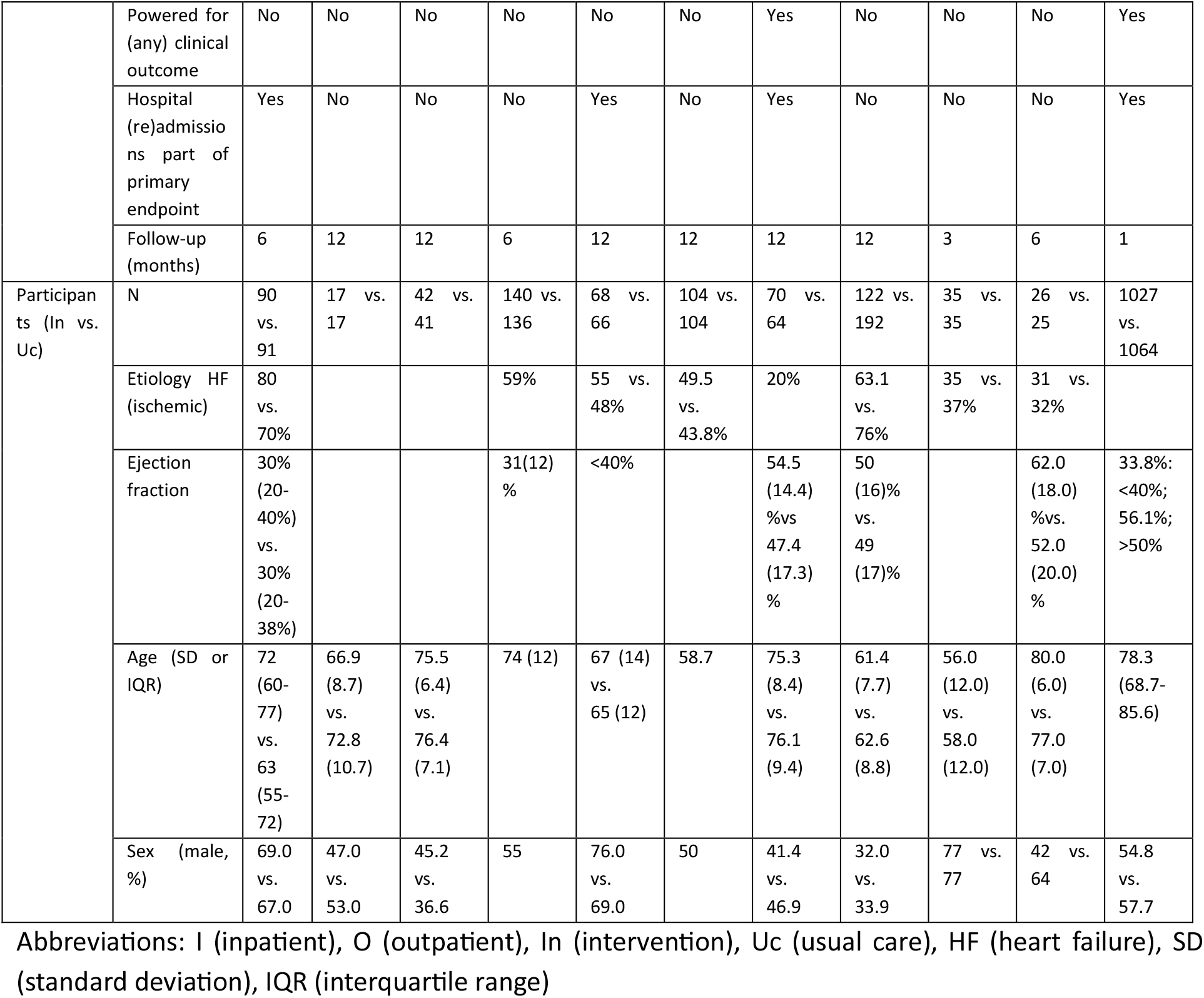
Included studies.

Demographic details of study participants showed considerable diversity. Ages spanned from 58 to 80 years, and the proportion of male participants ranged from 32 to 77%. The proportion of ischemic HF etiology was not reported in 3 studies^29,30,35^ and varied significantly, with the highest reported at 80%^28^ and the lowest at 20%^38^. Additionally, EF values displayed a wide range, reflecting a mix of HF cases with both reduced and preserved EF.

### Risk of study bias

Overall, the included studies demonstrated a low risk of bias across all domains (Figure 2). No high risk was identified for any of the included RCTs. Specifically, most trials exhibited low risk with regard to the randomization process, adherence to the intended intervention, completeness of outcome data, accurate outcome measurement, and appropriate selection of reported results. Some concerns were noted in a few studies, ^30,34,36,37^ mostly due to incomplete wording in the methods section on how the randomization process was implemented or how missing outcome data were managed. Despite these minor concerns, the overall assessment indicates that the studies generally maintained a robust methodological quality.

**Figure 2:**
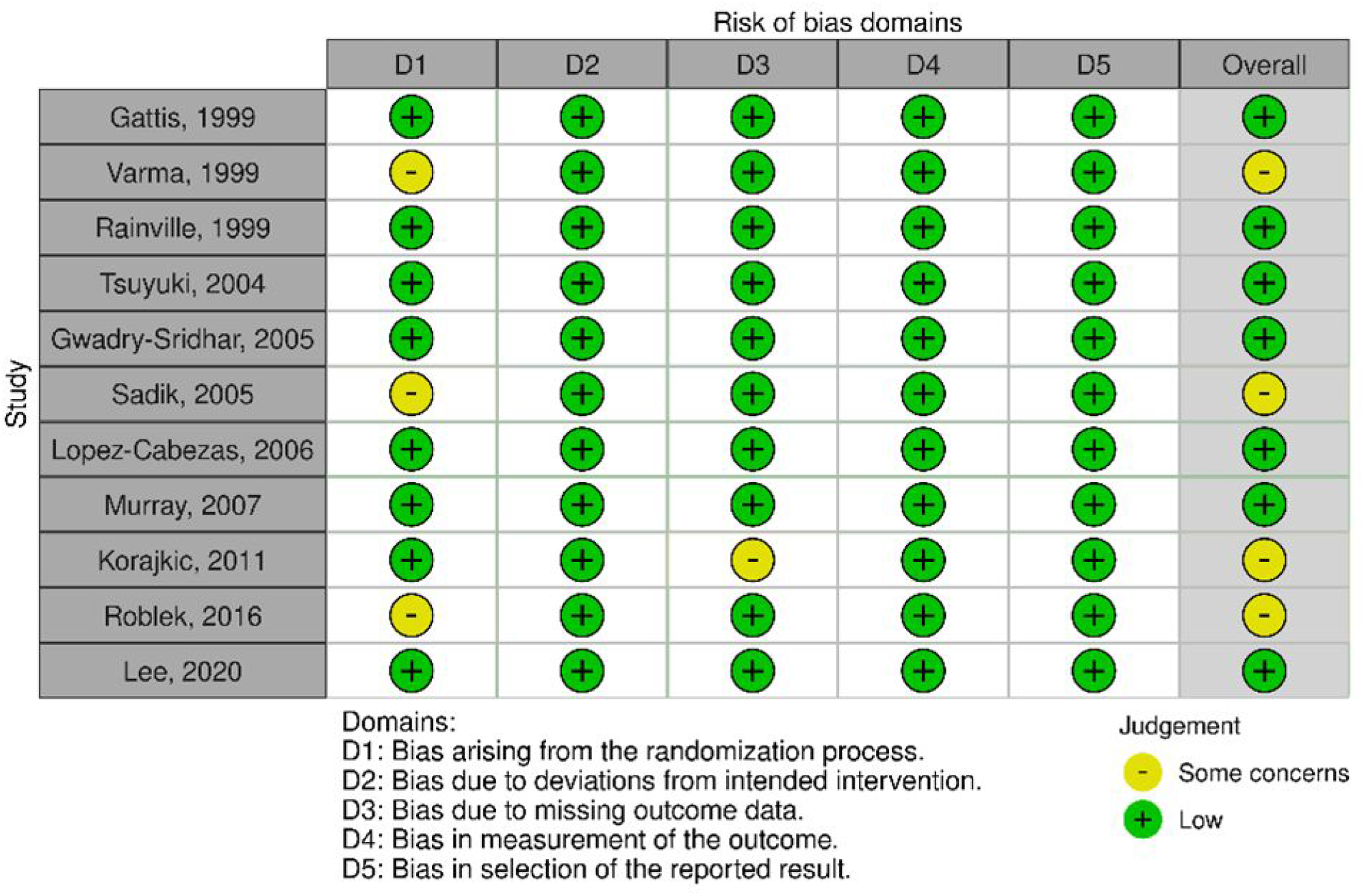
Risk of bias using the Cochrane RoB2 tool for the included randomized controlled trials.

### Meta-analytical results

Two separate meta-analyses were conducted, the results of which have been summarized in forest plots in Figure 3. In the Supplementary Appendix, the complementary drapery plots can be found for each meta-analysis, showing the p-value distribution per study (Figure S1 and Figure S2).

**Figure 3:**
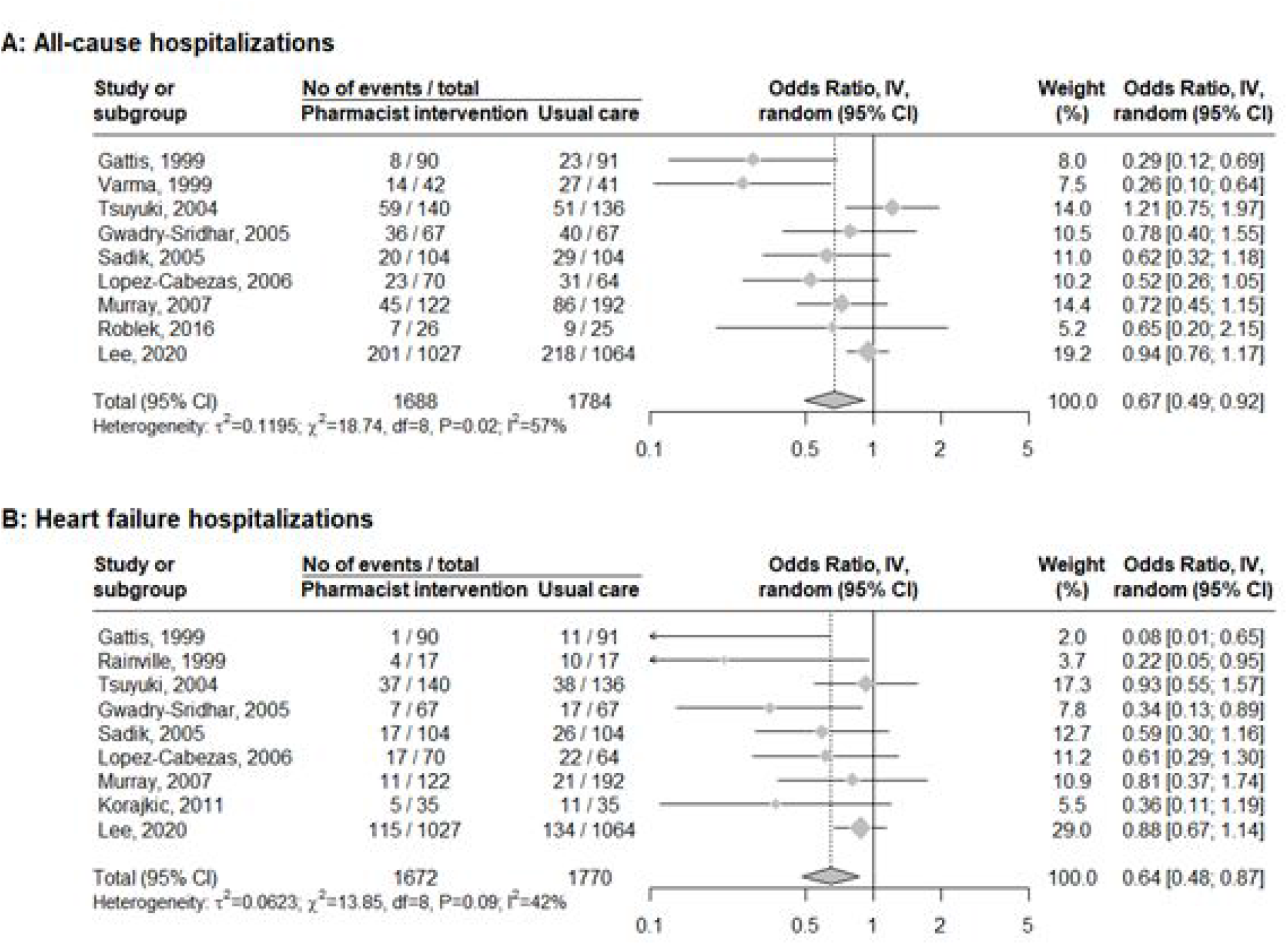
Forest plots from the meta-analysis comparing pharmacist interventions versus usual care in heart failure. Panel A shows all-cause hospitalizations and Panel B heart failure hospitalizations.

The meta-analysis for all-cause hospitalizations included 9 RCTs with a total of 3472 participants (n_intervention_= 1688, n_usual care_ = 1784 patients) and 927 events. The random effects model showed an OR of 0.67 (95% CI: 0.49–0.92, p=0.0119) in favor of pharmacist interventions. The Bayesian meta-analysis resulted in a comparable OR of 0.67 (95% credible interval: 0.44-0.98). Heterogeneity was moderate with an I^2^ of 57.3% (95% CI: 10.3–79.7%) and significant with a p-value of 0.016 (test of heterogeneity, Q=18.74). The NNT was estimated at 26 (95% CI: 16-103).

A total of 9 RCTs, totaling 3442 observations (n_intervention_=1672, n_usual care_=1770) and 504 events, were included in the meta-analysis with HF hospitalizations as outcome. The random effects model yielded an OR of 0.64 (95% CI: 0.48-0.87; p=0.0038). This translated into a NNT of 23 (95%: 16-65). The overall OR remained significant, also at the p<0.01 level, as shown in the drapery plot (Supplementary Appendix, Figure S2). The Bayesian estimate provided an OR of 0.61 (95% credible interval: 0.37-0.92). Heterogeneity analysis indicated moderate variability among studies (I^2^ = 42.2% (95% CI: 0.0-73.4%), p = 0.0858).

### Publication bias

The funnel plots for both all-cause and HF hospitalizations showed comparable asymmetry (Figure 4). There was a visual association between OR values and precision, with lower OR values mostly found in studies with lower precision, which was confirmed by Egger’s test (all-cause hospitalizations p=0.0297; HF hospitalizations: p=0.0013).

**Figure 4:**
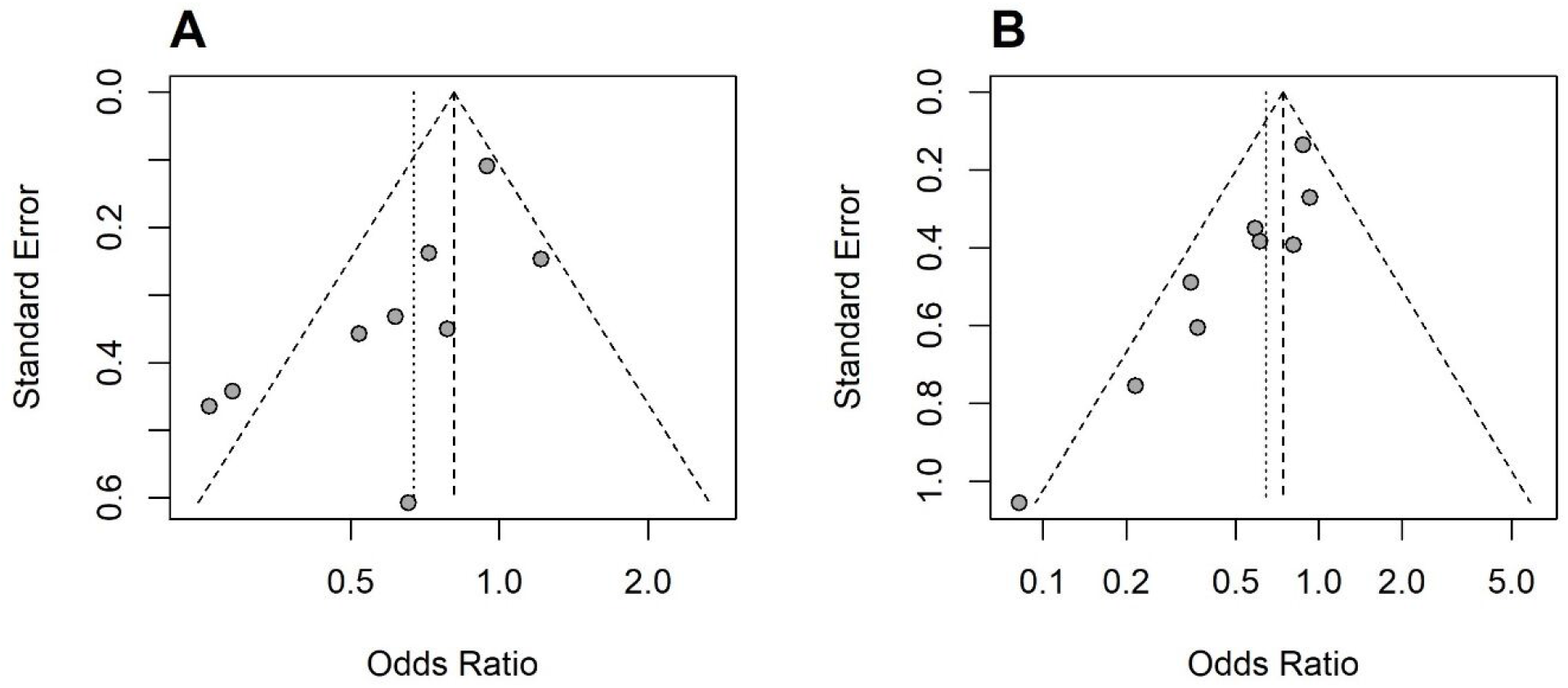
Funnel plots. Panel A (all-cause hospitalizations), panel B (heart failure hospitalizations).

### Sensitivity and subgroup analyses

The sensitivity analysis for all-cause hospitalizations did not identify a single study that changed the overall conclusion of the meta-analysis; the upper boundary of the 95 CI remained below 1 in the leave-one-out analysis (Figure 5A). A similar result was retrieved for HF hospitalizations (Figure 5B). Detailed results of both sensitivity analyses have been tabulated and added to the Supplementary Appendix (Table S2).

**Figure 5:**
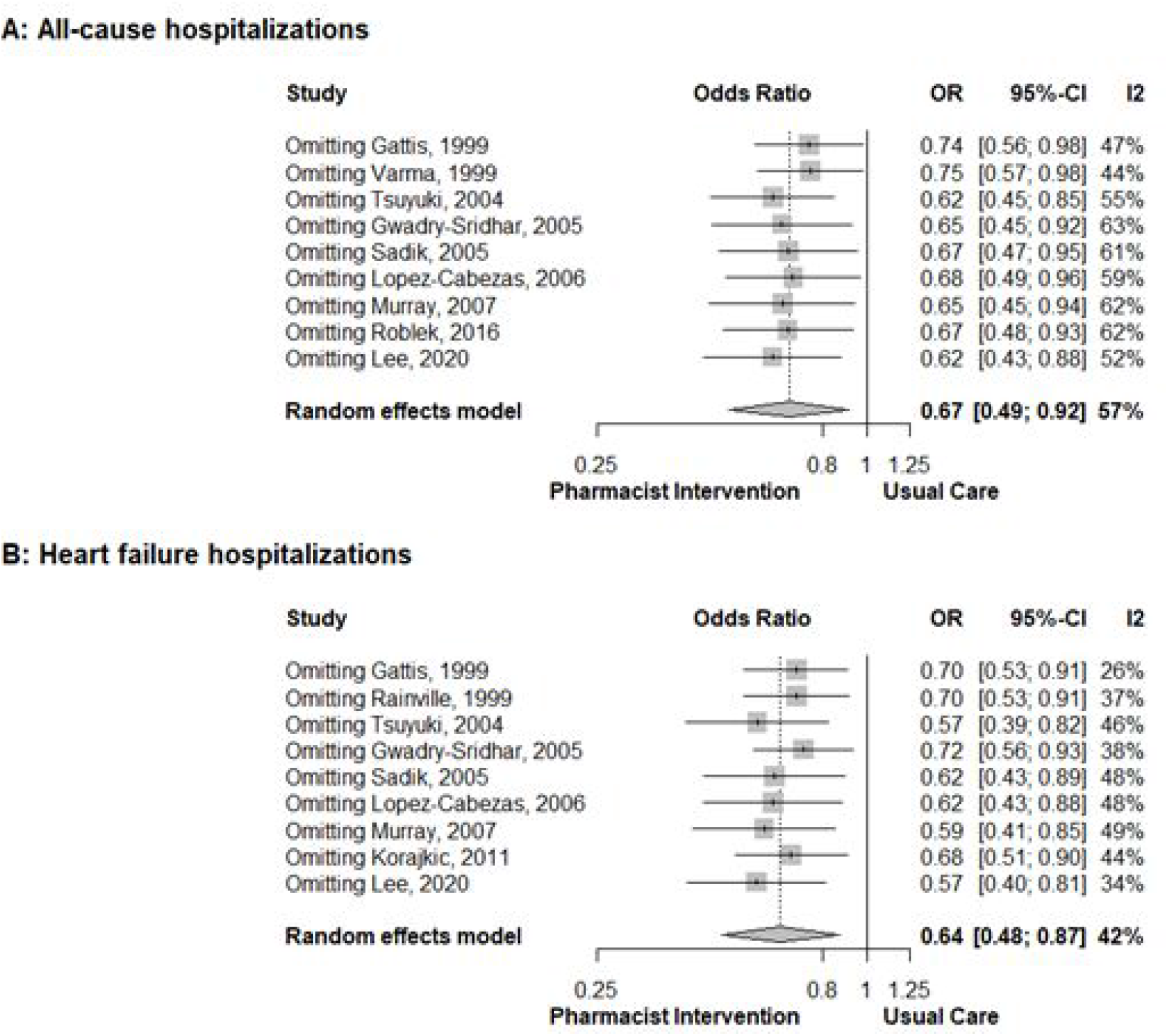
Forest plots of the leave-one-out meta-analyses. Panel A (all-cause hospitalizations). Panel B (heart failure hospitalizations).

Explorative subgroup analyses for all-cause hospitalizations showed larger effects when outpatients were recruited and when the follow-up time was 12 months. Non-significant trends suggested a larger reduction of these hospitalizations if adherence was promoted and education was provided. For HF hospitalizations, medication reconciliation, an extended intervention and the absence of sample size estimation were associated with lower OR values (Supplementary Appendix, Table S3).

## Discussion

Our systematic review and meta-analysis synthesized data from 11 RCTs, totaling 3576 patients (n_intervention_ = 1741, n_usual care_ = 1835) to evaluate the impact of health-system pharmacist interventions on hospitalizations in patients with HF. Both all-cause and HF hospitalizations were significantly reduced among in- and outpatients. The odds of hospitalizations were reduced by approximately one-third with relatively broad 95% CI boundaries. The robustness of our findings was confirmed by the leave-one-out analysis. For all-cause hospitalizations, the NNT was 26, which represents a clinically relevant effect size. This effect is plausible given that pharmacy interventions promote appropriate medication use and medication adherence in HF.^16^

Our findings have important implications for healthcare practice and HF management. The observed reduction in hospitalizations suggests not only improved patient outcomes but also potential cost savings. These effect sizes should encourage payers, hospital boards and medical teams to actively involve pharmacists in HF care.^39^ A recent review by Saverese *et al*. advocated for a team-based approach to improve HF care throughout the patient journey.^40^ Their proposal closely aligns with conclusions made nearly 25 years ago by Gattis *et al*. Indeed, Gattis *et al*. concluded that improving complex HF care over time necessitates a multidisciplinary team approach.^28^ Current clinical practice guidelines also recommend team-based care given the complexity of care in HF and the challenges of patient follow-up.^10,41^ Our analysis reaffirms the findings of the PHARM study by Gattis *et al*. and further supports the need for increased involvement of pharmacists in HF care. For pharmacists themselves, there is another—albeit less obvious—implication. Pharmacists are often stretched thin in hospital settings. Our data suggest that prioritizing HF within the responsibilities of hospital pharmacists may enhance their effectiveness and overall impact, particularly in comparison to other interventions.

Most studies in our review evaluated pharmacist interventions that included a variety of components, with several studies examining similar approaches.^28,30,37^ Many of the included studies were small, and underpowered to detect significant clinical effects. Since underpowered studies are more prone to overestimate their results, the conclusion of our meta-analysis should be interpreted with care. Additionally, only one of the studies was conducted in Europe, which might be explained by differences in healthcare systems and practices. In Europe, there is often a preference for staffing HF outpatient clinics with specialist nurses, rather than pharmacists. This might partly also be influenced by a reluctance among pharmacists to assert themselves in such clinical roles. Most RCTs assessed pharmacist interventions within a team environment that included other healthcare professionals, such as nurses. Jack *et al*. already pointed out that a nurse-pharmacist dyad is highly effective to reduce hospital admissions.^42^

Funnel plot asymmetry, confirmed by Egger’s test, indicated that smaller studies reported more positive results with a lower precision, potentially due to publication bias. It might also be explained by the inherent heterogeneity of the studied pharmacist interventions. Additionally, earlier, smaller trials from the late 90’s with positive findings may have contributed to the observed asymmetry.^28,29,43^ Importantly, our sensitivity analyses confirmed the stability of our findings, with no single study altering the conclusion that pharmacist interventions reduced hospitalizations.

The four previously highlighted meta-analyses published between 2008 and 2021, included varying numbers of RCTs and approached pharmacist involvement in HF care from different perspectives. Koshman *et al*. provided a first meta-analysis and incorporated 12 RCTs. They reported a significant reduction in both all-cause (OR 0.71, 95% CI 0.54-0.94, p=0.02) and HF hospitalizations (OR 0.69, 95% CI 0.51-0.94, p=0.02).^17^ In their update, Parajuli *et al*. included 16 RCTs and demonstrated significant reductions in all-cause (OR 0.76, 95% CI 0.60-0.96, p=0.02) and HF hospitalizations (OR 0.72, 95% CI 0.55-0.93, p=0.01).^18^ In 2021, Arunmanakul *et al*. analyzed 29 RCTs and found a more modest reduction in all-cause admissions (OR 0.87, 95% CI 0.77-0.99, p=0.041).^20^ In their analysis, they included a subgroup analysis of hospital-based settings with ambulatory care that showed a more substantial reduction of all-cause hospitalizations (OR 0.57, 95% CI 0.44-0.75). That same year, Schumacher *et al*. focused on outpatient, community and home-based pharmacist interventions across 24 RCTs. They did not find a statistically significant reduction in all-cause (OR 0.86, 95% CI 0.73-1.03, p=0.10) and HF hospitalizations (OR 0.89, 95% CI 0.77-1.02, p=0.10).^22^ In contrast, our analysis, which included 11 RCTs in total, revealed significant reductions in both all-cause (OR 0.67, 95% CI 0.49-0.92, p=0.0119) and HF hospitalizations (OR 0.64, 95% CI 0.48-0.87, p=0.0038) with comparable effect sizes. Notably, the data for the latter endpoint were convincing.

Our decision not to include mortality as a primary outcome in our meta-analysis was intentional. By focusing on hospitalizations, we were able to assess a more immediate and tangible outcome directly influenced by pharmacist interventions, providing clearer insights for healthcare resource utilization and cost-effectiveness. From a payer’s perspective, this measure is particularly relevant. In a recent analysis by Van Wilder *et al*., the authors assessed the variability of cardiovascular outcomes (as a surrogate for the quality of care) in a Belgian hospital cohort.^27^ They found that HF hospitalizations accounted for approximately 10% of all cardiovascular admissions, with 30-readmission rates at nearly 15% - a figure consistent with current literature and one that showed considerable variability between hospitals.^8^ The authors concluded that the quality of care in HF requires greater attention. Given that healthcare costs are significantly driven by these frequent hospitalizations, reducing this outcome by about one-third would substantially lower costs in a cost-effective manner. We argue that health-system pharmacists can and should play a role in reducing this variability of care.^39^

The strength of our study lies in the rigorous methodology applied to synthesize data from diverse RCTs spanning over two decades and various healthcare settings. However, our analysis was limited by the variability in intervention types and outcome measures among included studies, which may affect the generalizability of our findings to specific patient populations or healthcare contexts. Importantly, many RCTs were conducted in the U.S., which might limit their validity to the European healthcare setting. Moreover, the high heterogeneity between RCTs (i.e.., relatively high values of the I^2^ statistic) and the potential for study bias (i.e., significant Egger’s test) should caution us from overinterpreting the data. Additionally, even though most studies were methodologically sound, the majority were not powered to detect differences in clinical outcomes.

Moving forward, future research should focus on refining the implementation of pharmacist interventions, with the goal of integrating these strategies into routine clinical practice for HF management. Investigating additional patient-centered outcomes, such as quality of life, could provide a more comprehensive understanding of the broader impact of pharmacist-led interventions in cardiovascular care. Further research is also needed to explore how pharmacists can optimize GDMT over time, particularly within larger healthcare systems, such as through independent prescribing of GDMT.^14^ Currently, team-based care, with pharmacists as key members, is already recommended in both U.S. and European clinical practice guidelines on HF management, with a class IA recommendation, given the complexity of HF management across multiple care transitions.^10,41^ Finally, future studies should further explore the cost-effectiveness of these pharmacist interventions in specific regions, accounting for patient demographics and the unique characteristics of local health systems.

## Conclusion

Our systematic review and meta-analysis found that health-system pharmacist involvement in HF management reduced the risk of both all-cause and HF hospitalizations by about a third. Given the high hospitalization rates among in- and outpatients with HF, increasing the integration of health-system pharmacists into multidisciplinary care teams should be an absolute priority.

## Contributors

The concept was proposed by LVdL, who also conducted the analyses, created the tables and figures, and drafted the initial manuscript. LVdl, CG and PF screened and curated the data. All authors made substantial contributions to the interpretation of the data, reviewed the manuscript, and provided final approval of the submitted version.

## Funding

LVDL has received funding from the Clinical Research Fund of UZ Leuven. The funder had no active role in generating this report.

### Competing interests

LVdL, CB, CV, FK-C and LVA have no conflicts of interest to declare. PF has received previous honoraria for scientific advice, lecture fees, research, and/or educational grants from AstraZeneca, Novartis, Novo Nordisk, Pharmacosmos, Servier, and Vifor. RT has received investigator-initiated research grants from Merck, AstraZeneca, Pfizer, and Sanofi, and has consulted for Shoppers Drug Mart, Merck, and Emergent Biosolutions.

### Patient consent for publication

Not applicable.

### Ethics approval

Not applicable.

## Data availability statement

The datasets generated and analyzed during the study are available from the corresponding author on reasonable request.

## Notes

### Clinical Trial

N/a (no intervention trial was conducted as part of this manuscript)

### Author Declarations

not applicable, given the type of manuscript, ie a systematic review & meta-analysis.

